# Scoring Rubric to Assess Ethical Issues in Clinical Artificial Intelligence Tools

**DOI:** 10.64898/2026.09.05.26362340

**Authors:** Kevin Kindler, Shyam Visweswaran, Yanshan Wang, Jose Abad, John Maier, Olga Kravchenko

## Abstract

With the growing use of artificial intelligence (AI) tools in clinical care, it is essential to assess ethical issues related to using these tools, both before and after deployment. To address this need, a multidisciplinary team of clinical and AI experts developed and tested a scoring rubric to evaluate AI tools in clinical settings from an ethical perspective, considering potential issues that may arise during their use. The rubric provides a replicable scoring system comprising five content areas and four scoring categories. The content areas include evaluation population, accessibility, clinical bias, user control, and monitoring and updates. To test the applicability of the rubric, we applied it to three AI tools designed for clinical decision-making, clinical documentation, and clinical image analysis. The evaluation relied solely on publicly available information to simulate typical clinical settings. The rubric addresses a critical gap in the evaluation and governance of clinical AI tools by providing a structured framework to assess their ethical aspects. This approach enables healthcare systems to address and monitor the ethical impact of AI tools, fostering greater trust among clinicians and patients and guiding improvements in clinical practice.

## INTRODUCTION

As artificial intelligence (AI) tools increasingly permeate clinical care, it is critical to evaluate their ethical qualities, including respecting patient rights, promoting equitable care, and being free of bias, both before and after deployment. AI tools that lack transparency and generalizability, have clinical bias, exhibit barriers to accessibility, or inhibit appropriate governance can result in poor outcomes or increased disparities among patients. AI applications intended for clinicians can be broadly divided into four categories: diagnosis, clinical care, health monitoring, and healthcare administration.^1^ For example, diagnostic tools can analyze patient data or medical images for early signs of disease; clinical care solutions can formulate personalized treatment plans or guide surgical procedures; health monitoring systems can continuously track vital signs and alert the healthcare team about potential problems; and healthcare administration tools can streamline patient scheduling and billing. Further, AI tools intended for patients that help with diagnosis, clinical care, and health monitoring require a higher degree of ethical review.

The development and deployment of clinical AI tools is a multi-stage process, from initial design to deployment in the clinical setting, which involves rigorous evaluations at each stage to ensure safety, efficacy, and reliability. These evaluations encompass pre-deployment *in-silico* testing (e.g., simulation-based), silent (offline) evaluation, where the AI is tested behind the scenes without influencing clinician behavior or patient outcomes, early live clinical evaluation, prospective live clinical evaluation, and post-deployment ongoing monitoring. Several assessment rubrics and reporting guidelines have been developed, each appropriate to one or more stages. For example, TRIPOD-AI and STARD-AI are intended for pre-clinical evaluation, whereas DECIDE-AI, CONSORT-AI, and SPIRIT-AI focus on clinical evaluation.^23456^ However, no rubrics or guidelines are available for evaluating ethical issues in clinical AI technologies. An ethical rubric would enable healthcare organizations at all levels, ranging from the entire system to individual clinical departments, to objectively evaluate the presence of ethical concerns in an AI tool. Reproducible evaluation of an AI tool could build confidence among clinicians wishing to use it, as it provides generalizability across different healthcare systems. Additionally, having an objective, quantitative rubric for evaluating ethical concerns may increase patient trust.

We hypothesize that introducing a rubric to assess ethical issues in AI tools being considered for deployment or already in use will enable objective assessments of these technologies before and after deployment. This approach will encourage healthcare organizations to carefully examine emerging tools for ethical concerns, establish an objective scoring system for comparing tools, and foster greater trust among clinicians and patients.

## METHODS

### Rubric Development

The rubric was developed by a multidisciplinary faculty team (n=5) assembled from an institutional working group focused on ethical issues in clinical AI, including family medicine and neurology clinicians, informaticians, and AI experts. The rubric development process consisted of iterative rounds of discussions focused on refining content areas and scoring criteria, followed by applying the final rubric to three AI tools currently in clinical use.

The initial rubric content areas and scoring categories were identified from the existing literature and clinical practice needs. The initial rubric was then modified by adopting concepts from the Grading of Recommendations, Assessment, Development, and Evaluations (GRADE) framework, which is used for rating the quality of evidence and strength of recommendations for clinical practice guidelines.^10^ GRADE categorizes evidence quality into four levels: high, moderate, low, and very low, and it allows the upgrade or downgrade of these levels based on criteria such as risk of bias, imprecision, large dose effect, and others.^11^ While GRADE focuses on a single aspect (quality of evidence), we expanded this idea to scoring multiple content areas, replaced quantitative categories with numerical scores, and developed a composite score obtained by totaling the individual domain scores.

In the first round of discussion, three members of the interdisciplinary expert panel (KK, OK, JA) independently rated the importance of each content area in the rubric for inclusion using a scale of low, medium, or high importance, and provided free-text suggestions for changes to the content areas and the scoring categories. The content areas and the scoring categories were revised, guided by the quantitative ratings and the qualitative feedback. Content areas that received a majority of low ratings were removed from the rubric. The remaining content areas were revised based on the qualitative feedback. Content area descriptions were revised to improve clarity, conceptually overlapping areas were consolidated to reduce redundancy, and scoring categories were reworded to remove ambiguity.

In the second round, the revised rubric was reviewed by a 17-member institutional Working Group on AI Ethics in Primary Care consisting of clinicians, informaticians, AI experts, and graduate students. The content areas and the scoring categories were revised based on the feedback and suggestions for modifications from the working group. In the third round, the revised rubric was evaluated by five members of the interdisciplinary expert panel (KK, OK, JA, SV, and JM) to achieve a final consensus. This panel was comprised of family medicine and neurology clinicians, operational and academic clinical informaticians, and AI experts.

Consensus was defined as unanimous agreement among panel members on the final rating for each content area and scoring category. The rubric was finalized based on feedback from the third round of review.

### Application of the Rubric

To assess the applicability and utility of the rubric, we applied it to three different types of AI tools currently in use: 1) UTICalc, a clinical decision support tool that estimates the probability of urinary tract infections (UTIs) in pediatric patients aged 2 to 23 months; 2) Abridge, an administrative tool that transcribes and summarizes medical conversations from clinical encounters, and generates clinical notes, after visit summaries, and patient education materials;

3) Medihub Prostate, a radiology-focused software tool that analyzes prostate MRI images to identify cancerous areas in the prostate gland and visually present the results for the use by radiologists and urologists.^789^ The rubric was applied to these tools using only publicly available information to simulate real-world conditions under which clinical departments might evaluate AI tools without privileged access to proprietary development data. This approach was designed to reflect the rubric’s intended use as a practical evaluation tool for healthcare settings.

Neither the development of the rubric nor its use in the evaluation of existing AI tools involved human subjects research or protected health information, and the University of Pittsburgh’s Institutional Review Board determined that the study was exempt

## RESULTS

We first describe the quantitative rubric for evaluating ethical issues in clinical AI tools and then the results obtained from applying it to three AI tools.

### The Rubric

The rubric comprises five content areas and four scoring categories (Table 1). The content areas included the population evaluation (population on which the tool was evaluated), accessibility (considerations for potential barriers to adoption and use), clinical bias (risk of bias and potential harm to the patient), user control (opportunities for human oversight), and monitoring and updates (capability for ongoing evaluation and system-level feedback). The scoring categories comprised supportive data (score = 2), possible limitation (score = 1), theoretical risk (score = 0), and known harm (score = −1). The rubric included a checklist specifying the scoring criteria, similar to the TRIPOD-AI framework.^4^ The checklist is shown in Table 2 and provides a detailed description of requisite content to satisfy individual checklist items and facilitate scoring. Application of the rubric to an AI tool generates a numerical score (ranging from 2 to −1) for each of the five content areas, and the sum of the scores yields an overall score (ranging from 10 to −5).

**Table 1.** Scoring rubric to assess ethical issues in artificial intelligence tools.

| Content Area | Scoring Category (Score) |  |  |  |
| --- | --- | --- | --- | --- |
|  | Supportive Data (2) | Possible Limitation (1) | Theoretical Risk (0) | Known Harm (-1) |
| Evaluation Population | Evaluated on a large and representative population | Evaluated on a small and unrepresentative population | Inadequate information on evaluation population | Not evaluated on any population |
| Accessibility | Freely accessible, or offered at a cost and always covered by insurance | Accessible for a fee, and sometimes covered by insurance | Accessible for a fee, and not covered by insurance | Not accessible |
| Clinical Bias | No documented risk of bias in diagnosis, treatment, or disparities in care | Low documented risk of bias in diagnosis, treatment, or disparities in care | No or limited evaluation of bias in diagnosis, treatment, or disparities in care | No evaluation of bias and likely risk of bias in diagnosis, treatment, or disparities in care |
| User Control | User has complete ability to modify or override the tool's output, or influence its behavior | User has partial ability to modify or override the tool's output, or influence its behavior | User has no ability to modify or override the tool's output, or influence its behavior | User has no information on ability to modify or override the tool's output, or influence its behavior |
| Monitoring and Updates | Continuous monitoring of performance and frequent updates | Periodic monitoring of performance and infrequent updates | As needed monitoring and updates in response to a concern | No monitoring of performance and no updates |

| Content Area | Description |
| --- | --- |
| Evaluation Population | The evaluation population refers to the group of patients or individuals on whom the AI tool is tested, and it should be representative of the intended population. |
| Accessibility | Accessibility refers to the ease and affordability with which clinicians and institutions can access and effectively use the tool. |
| Clinical Bias | Clinical bias refers to a tool's systematic errors in specific populations caused by flaws in data, algorithms, or implementation, resulting in suboptimal care in those populations. |
| User Control | User control refers to the ability of clinicians to modify or override the tool's outputs and influence its behavior in order to provide safe, effective, and relevant patient care. |
| Monitoring and Updates | Monitoring and updates refer to the ongoing processes of evaluating the tool's performance, safety, and efficacy in real-world clinical settings, as well as making improvements or modifications. |

**Table 2.**
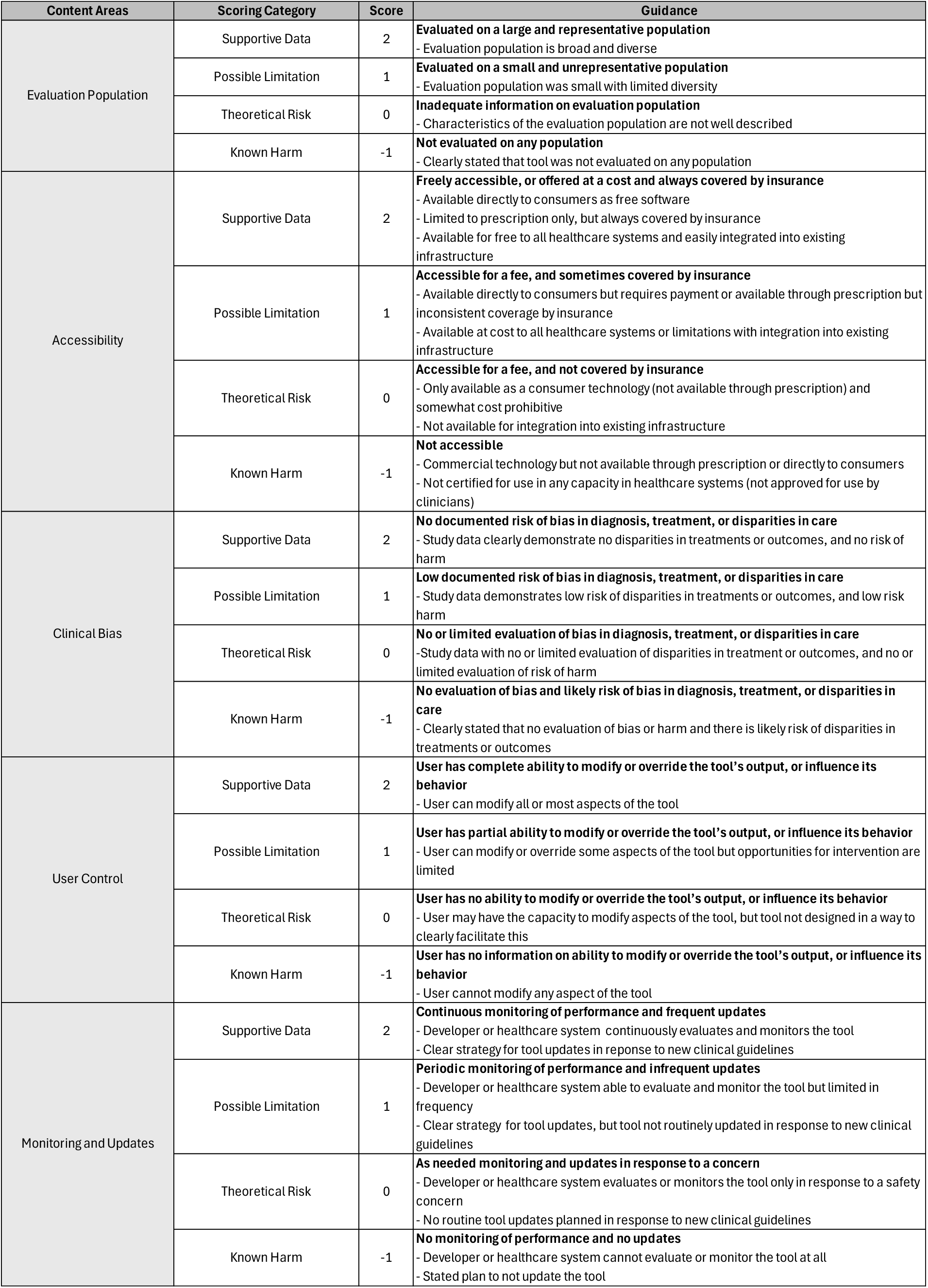
Detailed checklist of the information necessary to satisfy scoring categories within each content area for ethical concerns in clinical artificial intelligence.

### Evaluation of AI Tools

Applying the rubric to three AI tools resulted in the following total scores: UTICalc scored 6, Abridge scored 8, and Medihub Prostate scored 3 (Table 3).

**Table 3.**
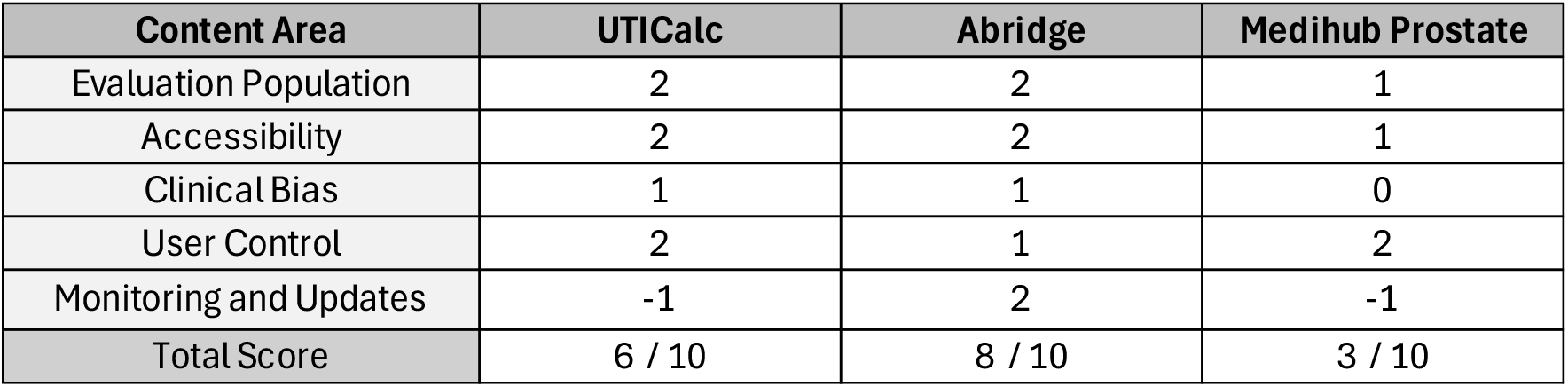
Scoring of three currently available clinical artificial intelligence tools.

| Content Area | UTICalc | Abridge | Medihub Prostate |
| --- | --- | --- | --- |
| Evaluation Population | 2 | 2 | 1 |
| Accessibility | 2 | 2 | 1 |
| Clinical Bias | 1 | 1 | 0 |
| User Control | 2 | 1 | 2 |
| Monitoring and Updates | -1 | 2 | -1 |
| Total Score | 6 / 10 | 8 / 10 | 3 / 10 |

UTICalc is on its third iteration and is available as a free web application.^8^ The current iteration of UTICalc was improved to mitigate racial bias.^12^ Points were deducted for clinical bias due to the possibility of harm (method of urinary collection in a child), the inability to monitor performance from a health system perspective, and the lack of information on updates to the tool. Abridge’s tool is available at a cost to healthcare systems.^9^ Points were deducted because of the risk of harm if the output documentation incorrectly reflects an aspect of a conversation and the limited ability to change the tool’s behavior if it makes errors in transcribing a conversation. Medihub Prostate was recently certified by the Food and Drug Administration (FDA) and has less supporting information than other tools.^10^ The evaluation data provided in the FDA documentation revealed a very small, narrow patient population with little demographic variability. The tool is available via a web portal, but organizational support is required to use the tool. Given the study’s small sample size, there is a potential for harm (misinterpretation of imaging studies used to diagnose prostate cancer) and treatment disparities. While the tool allows manual adjustment of all parameters, a healthcare system has no ability to assess the tool’s performance.

## DISCUSSION

A multidisciplinary group of clinical and AI experts developed a scoring rubric for ethically evaluating clinical AI tools, comprising assessments in five ethically relevant areas. The rubric was applied to three AI tools in clinical use, and the scores reflected areas of strength and weakness. The clearly defined criteria shown in Table 2 enable replicable AI tool analysis both within and across healthcare systems. The rubric enables rapid reevaluation of a tool should the tool be updated or additional information about testing protocols, plans for updates, and other important information becomes publicly available. The white paper from Abridge is an example that provides a clearly defined update release schedule, post-deployment monitoring, and validation practices.^13^

This rubric addresses a known gap in the literature and offers a practical solution for evaluating the ethical aspects of AI tools that are being considered for deployment or are already in use in healthcare. While each healthcare organization has its protocols for managing a digital portfolio, the clarity and simplicity of the rubric make it suitable for use in any healthcare system within an existing governance framework. Since this rubric addresses primarily ethical issues related to AI, organizations must conduct their due diligence to ensure that all other areas, such as accuracy, security, value, and scalability, are adequately addressed. Many of the content areas not covered by this rubric might be addressed using the Assurance Standards Guide created by the Coalition for Health AI (CHAI).^14^

We found that evaluating distinct AI tools demonstrated the rubric’s broad applicability. The assessment of the tools highlighted the value of incorporating perspectives from both clinical and AI experts. Thinking through the potential clinical implications of the output of a tool is just as important in evaluating a tool as understanding the elements of the assessment rubric. The three tools that were assessed relied on input from all content experts to conduct the evaluations. Future work will expand the application of the rubric to additional content areas relevant to AI, as well as discussions with healthcare systems regarding assessments of their AI portfolio.

## Data Availability

All data produced in the present work are contained in the manuscript

## ACKNOWLEDGMENTS

We thank the members of the Working Group on AI Ethics in Primary Care at the University of Pittsburgh for their invaluable input and collaboration.

## Notes

### Competing Interest Statement

The authors have declared no competing interest.

